# Patient Experiences with Diagnostic Revision in Lymphoma: Analysis of Chinese Online Forum Narratives Using Natural Language Processing

**DOI:** 10.1101/2025.09.07.25335273

**Authors:** Feng He, Zecheng Yang, Xinyu Zhang, Wee Ling Koh, Loraine Liping Seng, Nicholas Goodwin, Jose M Valderas

## Abstract

**Background:** Lymphoma diagnosis is challenging due to diverse subtypes and nonspecific presentations, with high rates of revision documented across settings. Patient experiences with diagnostic revision remain poorly understood, yet they can reveal gaps in diagnostic pathways. We analysed lymphoma forum narratives to characterise patient experiences during diagnostic revision in terms of barriers, facilitators, and the associated burden.

**Methods:** This cross-sectional study used publicly accessible threads from China’s largest lymphoma forum (House086.com, 2011-2025). Narratives involving diagnostic revision were identified and extracted into structured case profiles using DeepSeek-V3.2 large language model and optical character recognition. DeepSeek assigned each case to either mild-to-moderate or severe burden status, with classification reliability assessed through rubric-guided human audit. Separately, transformer-based keyword clustering identified eleven barriers and seven facilitators along the diagnostic revision pathway. Associations between barriers/facilitators and severe-burden status were examined using multivariable logistic regression.

**Results:** We identified 1,801 unique cases involving diagnostic revision (median age 46, 58.4% family-authored). DeepSeek assigned 40.3% to severe-burden status, with human audit showing substantial agreement (Cohen’s κ=0.64, n=350). The most frequently mentioned facilitators were specialist input, tertiary hospitals, and patient self-advocacy (all >50%). The most common barriers were clinician-related issues (e.g., inadequate communication; 90.4%) and case complexity (74.1%). Severe-burden status was associated with all eleven barriers, with the strongest association observed for prior inappropriate treatment despite its low frequency (9.2%; OR 37.76, 95% CI 21.59-66.04). Among facilitators, clinician expertise (11.8%; OR 0.34) and peer networks (18.8%; OR 0.49) showed the strongest associations with lower odds of severe-burden status.

**Conclusion:** In narratives reporting lymphoma diagnostic revision, burden severity was associated with factors across clinical, system, and patient levels. Peer networks emerged as a meaningful source of support complementing formal care. Less frequent but high-risk events such as prior inappropriate treatment warrant priority attention.

**Funding:** Institutional support from CRiHSP.

## INTRODUCTION

Lymphoma accounts for 3-4% of all cancers worldwide, with more than half a million new cases reported annually.^1^ Accurate diagnosis remains a challenge due to diverse subtypes and nonspecific symptoms. Studies from the United States, United Kingdom, and Europe reported diagnostic revision in 5.8-27.3% of referred cases following expert pathological reviews, with revision rates as high as 39.8% in China.^2-5^ Uncertainty in diagnosis can delay appropriate care, expose patients to harmful treatments, and lead to long-term health consequences.^2,6^ Prior efforts have primarily focused on improving accuracy through clinician training, advanced imaging, or pathology support.^2,3,5-8^ Yet little is known about how patients interpret and respond to diagnostic changes. In many cases, revising an initial diagnosis requires patients and families to advocate for themselves, seek second opinions, and navigate financial and psychological burdens.^6,9^ The 2024 Global Patient Survey (GPS) by the Lymphoma Coalition, covering 11,170 respondents, highlighted persistent gaps in communication, shared decision-making, and psychosocial support across countries, ^10^ underscoring the need for more patient-centred lymphoma care.

Patient-reported experiences may help identify where breakdowns occur in the diagnostic pathway and highlight priorities for quality improvement, yet conventional surveys and registries capture these accounts only with limited depth and nuance.^11-13^ Patient narratives, with their vast scale and richness, offer a complementary perspective to existing literature despite lacking clinical verification.^11,13^ Existing methodologies for narrative analysis rely heavily on manual coding or keyword matching based on pre-defined categories,^12,14^ limiting scalability, semantic precision, and structural consistency. Recent advances in artificial intelligence (AI), particularly large language models (LLMs), enable large-scale analyses of unstructured, patient-generated data from online platforms.^14-16^ Applying AI to such data may help identify barriers, facilitators, and opportunities for diagnostic quality improvement.^16^

In this study, we aimed to: (1) identify barriers and facilitators relevant to diagnostic revision in lymphoma; (2) quantify their associations with the severity of burden; and (3) demonstrate the feasibility of analysing large-scale forum narratives using an AI-assisted approach.

## METHODS

### Study Design and Data Collection

We conducted a retrospective cross-sectional study of publicly accessible discussion threads from House086.com, the largest online forum dedicated to lymphoma patients and caregivers in China.^14^ Given the scale and heterogeneity of the data, this study employed an AI-assisted content analysis approach.^17,18^ Through an iterative exploratory process, we first compiled a list of keywords covering diverse topics relevant to diagnostic revision (Supplementary Table S1). We then manually screened all threads containing these keywords to curate candidate threads for extraction by our custom AI pipeline (Figure 1). Many threads included user-uploaded clinical materials, such as pathology reports, imaging reports, and hospital documents. We therefore applied optical character recognition (OCR) ^19^ to extract text from embedded images. The OCR-extracted text and discussion threads were then processed using DeepSeek-chat (V3.2, temperature=0.1), an LLM with strong performance in Chinese text comprehension.^20^ Each thread was processed through an independent stateless API call using the same prompt template.

**Figure 1.**
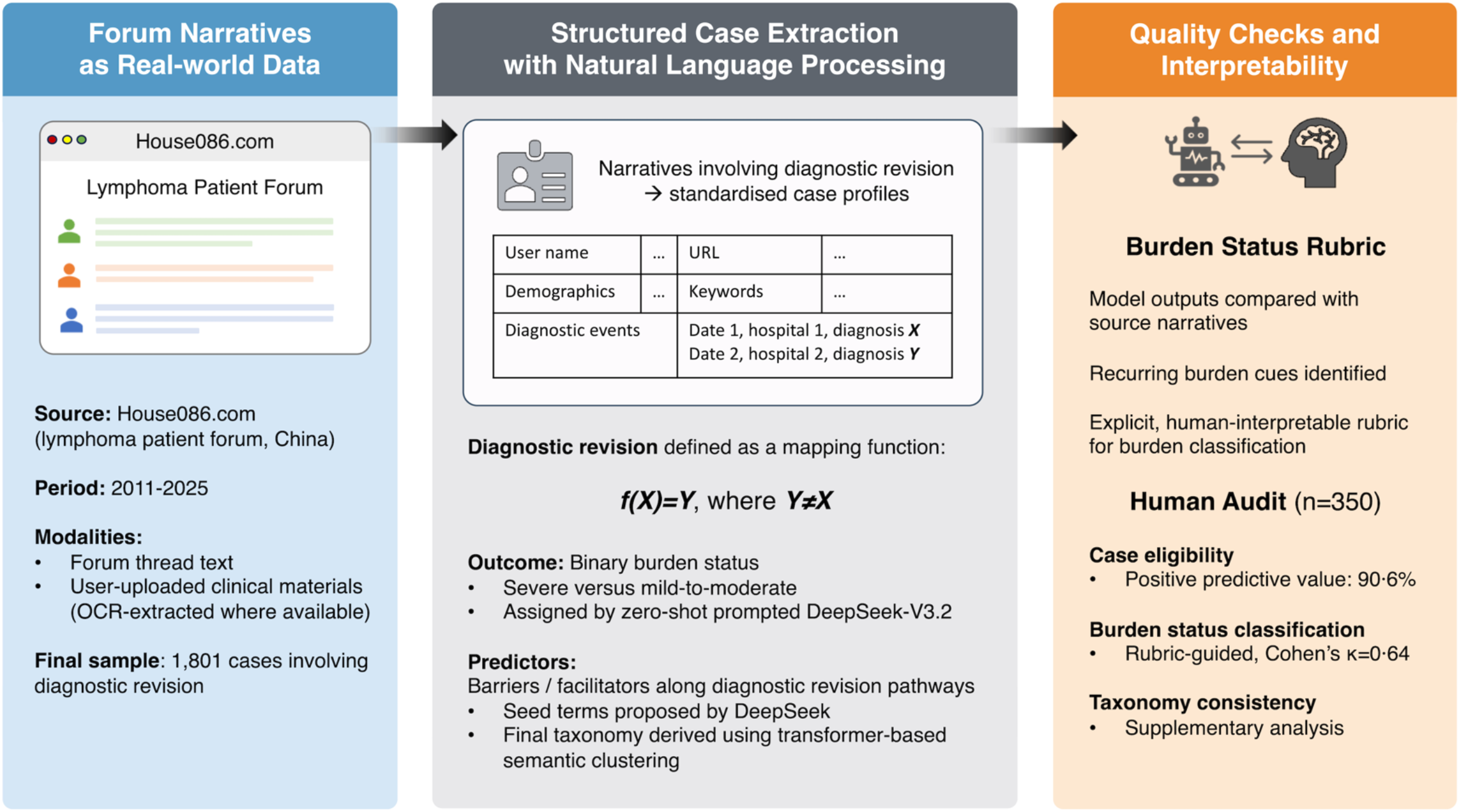
Pipeline for identifying, structuring, and auditing diagnostic revision cases from forum narratives. Publicly available forum narratives were transformed into structured case profiles using DeepSeek-V3.2 large language model, optical character recognition (OCR), and transformer-based keyword clustering. Diagnostic revision was operationally defined as a mapping from diagnosis X to Y (Y≠X). Study outcome was mild-to-moderate versus severe burden status assigned by DeepSeek. A human-readable rubric was developed to assess model interpretability and guide human audit.

### Prompt Engineering and Variable Definition

To mitigate the ambiguity in natural language, we defined diagnostic revision as a mapping function *f* from diagnosis *X* to diagnosis *Y*, with *Y≠X*. Specifically, our prompt instructed DeepSeek to identify narratives describing a completed or strongly implied revision path *f(X)=Y*, with eligible narratives required to contain details of *f, X*, and *Y* (Supplementary Method 1). Cases where *Y* was stated without a complete *f(X)=Y* trajectory were considered borderline, and reviewed manually for eligibility. These included cases of discordant pathological interpretations (e.g., both *X* and *Y* were reported without a definitive resolution *f*) or patient-reported misdiagnosis (e.g., narratives focused on the revision process *f* and outcome *Y* while omitting details of *X*).

For each identified narrative, the pipeline structured the corresponding contents into a standard case profile. Username, narrator type (patient, family member, or others), and year of posting were directly parsed from the website, whereas patient age, sex, diagnostic events (date, hospital, diagnosis), and keywords describing barriers/facilitators of diagnostic revision were extracted by DeepSeek based on narrative content.

A score (1-5) reflecting the severity of the associated burden (e.g., time loss, psychological distress, financial strain) was assigned by DeepSeek via zero-shot prompting. To ensure interpretability, the model’s implicit scoring logic was reverse-engineered to develop an explicit human-readable rubric, through iterative comparison between LLM outputs and source narratives (Supplementary Method 2). Due to the concentration of scores in the 3-4 range, burden severity scores were dichotomised into mild-to-moderate (≤3) versus severe (score ≥4) burden status, which served as the binary outcome for primary analysis. Each case also received an LLM-graded confidence score (1-5) to reflect overall case extraction quality.

### Keyword Taxonomy and Clustering

Given the heterogeneity in narrative content and style, DeepSeek was first prompted to propose free-text keywords for salient barriers and facilitators along diagnostic revision pathways. These seed terms were predominantly Chinese colloquial expressions, supplemented by English medical abbreviations. To capture contextual and cross-lingual semantic similarity, we used a multilingual sentence-transformer model (paraphrase-multilingual-MiniLM-L12-v2) to generate word embeddings, then applied agglomerative hierarchical clustering with Ward linkage to group semantically related terms. Based on clinical literature and pilot results, the resulting clusters were manually reviewed and further consolidated into a taxonomy of eleven barriers (e.g., case complexity, specimen issues) and seven facilitators (e.g., clinician expertise, peer networks). In formal analyses, DeepSeek assigned up to three barriers and up to three facilitators per case profile, constrained to this taxonomy.

### Data Preprocessing

The inclusion and exclusion workflow is summarized in Figure 2. From more than three million forum threads, 16,853 were returned by keyword search. Following manual review, 14,119 threads irrelevant to the study scope or duplicates were excluded. The remaining 2,734 unique threads underwent AI processing, of which 444 threads contained no eligible narrative, whereas the remaining 2,290 threads contributed 2,603 standardized case profiles related to diagnostic revisions. Of these 2,603 profiles, we excluded cases lacking meaningful description of diagnosis *Y* (n=111), and borderline cases deemed irrelevant upon manual review (n=322). For multiple narratives from the same user, only the most comprehensive and highest-quality version was retained (excluding n=361). Cases of low quality (confidence score ≤2, n=3) or missing LLM-graded severity scores (n=5) were also excluded. The final dataset comprised 1,801 unique user-level case profiles. Due to the anonymous nature of the online platforms, missing values in patient age and sex were observed. These values were all coded as “Not Disclosed” to retain sample size and preserve narrative diversity, with age categorised into consecutive groups for analysis.

**Figure 2.**
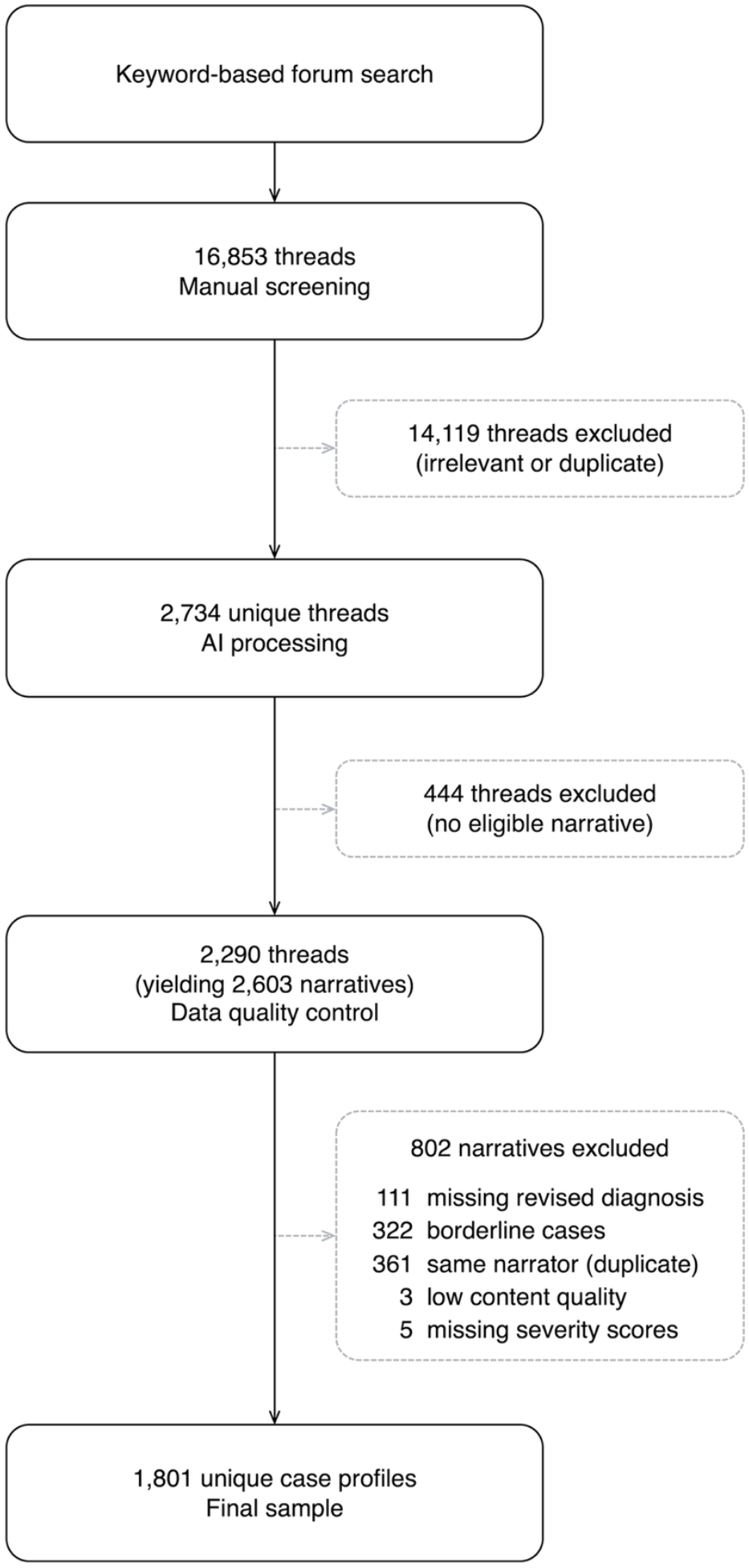
Data cleaning workflow. From more than three million forum threads, threads involving diagnostic revision were identified through keyword-based screening and manual review. Candidate threads were then processed through the NLP pipeline to identify relevant narratives and to extract structured case profiles. After quality control and de-duplication, 1,801 unique user-level case profiles were retained for analysis.

### Human Audit

To assess the interpretability of pipeline outputs, a stratified random sample of 350 cases was reviewed by a panel of biomedical professionals, with each case assessed by at least two reviewers. All reviewers were native Chinese speakers, with backgrounds spanning immunology, biostatistics, social work, and health services research. By examining the original forum contents blinded to pipeline outputs, reviewers assessed (i) whether a diagnostic revision was reported and (ii) the severity of its associated burden from the patient perspective, following the predefined rubric (Supplementary Method 2). Disagreements were resolved through reviewer discussion or majority vote where needed. The resulting consensus labels served as the reference standard for this audit.

Given the complexity and emotional intensity of patient narratives, reviewers were permitted to use LLM tools to assist with information extraction and clarification of clinical terminology.^21,22^ Nonetheless, final labelling decisions were made solely by human reviewers after verification against the original forum threads, including discussion texts, user profiles, and any uploaded clinical materials, to minimise potential AI hallucination or semantic drift. A supplementary audit was conducted to evaluate taxonomy consistency and potential extraction errors.

### Statistical Analyses

Continuous variables were summarized as medians with interquartile ranges (IQRs), and categorical variables as counts (%). Between-group differences were measured with the Mann-Whitney U test, or chi-square test as appropriate. To examine the associations between barriers/facilitators and severe-burden status in narratives involving diagnostic revision, we developed multivariable logistic regression models with the outcome defined above, adjusting for patient age, sex, narrator type, year of posting, and the number of diagnostic events. All numeric variables were categorized to improve model stability and accommodate potentially non-linear effects (Table 1). Pairwise associations among predictors were assessed using Cramér’s V to evaluate potential multicollinearity.^23^ Results were reported as odds ratios (ORs) with 95% confidence intervals (CIs), and p-values. To evaluate pipeline outputs in the human audit, case inclusion accuracy was calculated as the positive predictive value (PPV) of AI classification against human consensus labels. Human-AI agreement for assigning burden status (severe vs mild-to-moderate) was assessed using Cohen’s κ.

**Table 1.** Characteristics of narrative-derived cases by burden status.

|  | Overall<br>N=1801 | Mild-to-moderate<br>score≤3, n=1075 | Severe<br>score≥4, n=726 | p-value |
| --- | --- | --- | --- | --- |
| Patient age, years | 46.0 [29.0, 58.0] | 47.0 [31.0, 59.0] | 44.0 [28.0, 56.0] | 0.043 |
| Patient age category |  |  |  | 0.47 |
| <18 | 50 (2.8) | 26 (2.4) | 24 (3.3) |  |
| 18-40 | 244 (13.5) | 142 (13.2) | 102 (14.0) |  |
| 40-60 | 220 (12.2) | 129 (12.0) | 91 (12.5) |  |
| ≥60 | 150 (8.3) | 98 (9.1) | 52 (7.2) |  |
| Not disclosed | 1137 (63.1) | 680 (63.3) | 457 (62.9) |  |
| Patient sex |  |  |  | <0.0001 |
| Female | 527 (29.3) | 292 (27.2) | 235 (32.4) |  |
| Male | 661 (36.7) | 367 (34.1) | 294 (40.5) |  |
| Not disclosed | 613 (34.0) | 416 (38.7) | 197 (27.1) |  |
| Narrator type |  |  |  | 0.00030 |
| Patient-self | 709 (39.4) | 462 (43.0) | 247 (34.0) |  |
| Family member | 1051 (58.4) | 594 (55.3) | 457 (62.9) |  |
| Others (admin, doctors) | 41 (2.3) | 19 (1.8) | 22 (3.0) |  |
| Year of posting |  |  |  | <0.0001 |
| 2011 – 2015 | 298 (16.5) | 138 (12.8) | 160 (22.0) |  |
| 2016 – 2020 | 700 (38.9) | 412 (38.3) | 288 (39.7) |  |
| 2021 – 2025 | 803 (44.6) | 525 (48.8) | 278 (38.3) |  |
| No. of diagnostic events |  |  |  | 0.00020 |
| 2 | 1184 (65.7) | 744 (69.2) | 440 (60.6) |  |
| ≥ 3 | 617 (34.3) | 331 (30.8) | 286 (39.4) |  |

Continuous variables presented as median [IQR]; categorical variables as n (%). p-values from Mann-Whitney U test (continuous) or chi-square test (categorical). "Not disclosed" indicates information not available from the narrative.

All analyses were performed using R (version 4.2.0) and Python (version 3.11.13). Code and audit protocols will be made publicly available upon publication.

### Ethics

This study analysed patient narratives from an open-access online forum in China (House086.com). All data were publicly accessible without registration or login requirements, and the platform terms of use permit analysis of forum contents without further consent. No identifiable personal information was collected for analysis. Only de-identified and aggregate results are reported. The study protocol was reviewed by the National University of Singapore Institutional Review Board and was granted exemption from full review (NUS-IRB Reference Code: NUS-IRB-2025-1006). The study adheres to the principles of the Declaration of Helsinki.

### Role of the Funding Source

The study received institutional support from the Centre for Research in Health Systems Performance (CRiHSP), Yong Loo Lin School of Medicine, National University of Singapore. The authors had full responsibility for the study design, data collection, analysis, interpretation, and the decision to submit for publication.

## RESULTS

### Population Characteristics

Of the 1,801 cases identified from the lymphoma forum, 40.3% (726) were assigned to severe-burden status (LLM-graded score ≥4 on the 1-5 scale), whereas 59.7% (1,075) were assigned to mild-to-moderate burden status (≤3). Most cases were reported by family members (58.4%), followed by patients themselves (39.4%). A high proportion of cases did not disclose patient age (63.1%) or sex (34.0%). Family-authored cases were more frequently classified as severe-burden status than patient-authored cases (43.5% [457/1,051] versus 34.8% [247/709]). Cases involving multiple diagnostic revisions (≥3 diagnostic events) also had a higher proportion classified as severe-burden status than those involving a single revision (2 diagnostic events) (46.4% [286/617] versus 37.2% [440/1,184]). Over the study period (2011-2025), the number of identified cases increased by 169%, while the proportion with severe-burden status declined from 53.7% (160/298) in 2011-2015, to 41.1% (288/700) in 2016-2020, and 34.6% (278/803) in 2021-2025.

### Keyword Taxonomy and Frequencies

Transformer-assisted keyword clustering identified eleven barriers and seven facilitators for lymphoma diagnostic revision (Table 2). The most commonly mentioned facilitators were specialist input (e.g., external pathology review, 60.3%), tertiary-hospital care (e.g., nationally recognised centres with advanced infrastructure, 56.7%), and patient self-advocacy (52.7%) (Table S2). Among barriers, clinician-related issues (e.g., inadequate communication, 90.4%) and case complexity (e.g., pathological ambiguity, 74.1%) were most frequent. Three patient-level barriers (patient psychological resistance, family/financial constraints, and physical limitations) collectively appeared in 87 cases (4.8%) (Table S2). Given small cell sizes, these three low-frequency barriers were combined into a single composite variable to improve model stability for subsequent analyses.

**Table 2.**
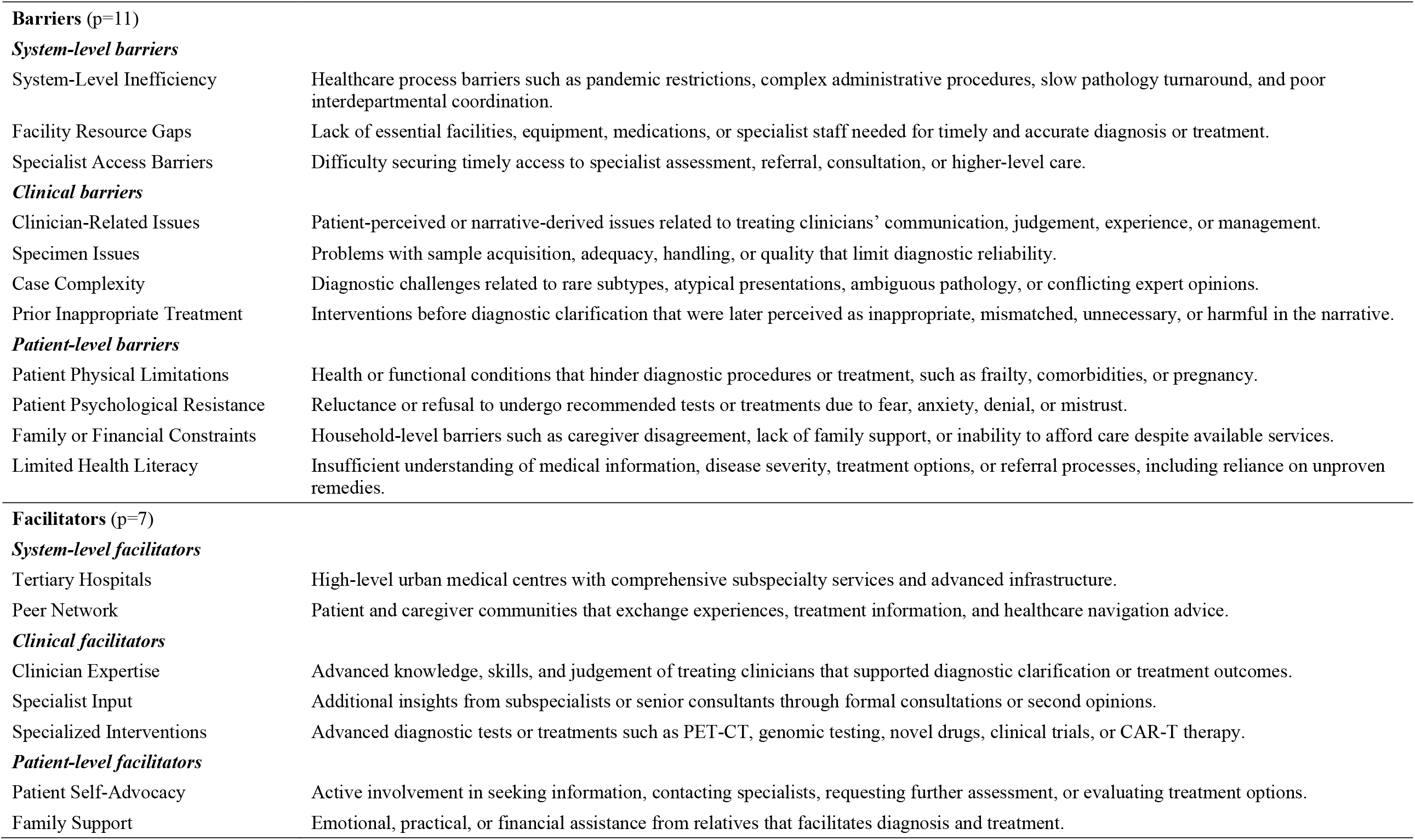
Narrative-derived taxonomy of barriers and facilitators in diagnostic revision pathways.

|  |  |
| --- | --- |
| System-Level Inefficiency | Healthcare process barriers such as pandemic restrictions, complex administrative procedures, slow pathology turnaround, and poor interdepartmental coordination. |
| Facility Resource Gaps | Lack of essential facilities, equipment, medications, or specialist staff needed for timely and accurate diagnosis or treatment. |
| Specialist Access Barriers | Difficulty securing timely access to specialist assessment, referral, consultation, or higher-level care. |
| ***Clinical barriers*** |  |
| Clinician-Related Issues | Patient-perceived or narrative-derived issues related to treating clinicians' communication, judgement, experience, or management. |
| Specimen Issues | Problems with sample acquisition, adequacy, handling, or quality that limit diagnostic reliability. |
| Case Complexity | Diagnostic challenges related to rare subtypes, atypical presentations, ambiguous pathology, or conflicting expert opinions. |
| Prior Inappropriate Treatment | Interventions before diagnostic clarification that were later perceived as inappropriate, mismatched, unnecessary, or harmful in the narrative. |
| ***Patient-level barriers*** |  |
| Patient Physical Limitations | Health or functional conditions that hinder diagnostic procedures or treatment, such as frailty, comorbidities, or pregnancy. |
| Patient Psychological Resistance | Reluctance or refusal to undergo recommended tests or treatments due to fear, anxiety, denial, or mistrust. |
| Family or Financial Constraints | Household-level barriers such as caregiver disagreement, lack of family support, or inability to afford care despite available services. |
| Limited Health Literacy | Insufficient understanding of medical information, disease severity, treatment options, or referral processes, including reliance on unproven remedies. |
| **Facilitators (p=7)*****System-level facilitators*** |  |
| Tertiary Hospitals | High-level urban medical centres with comprehensive subspecialty services and advanced infrastructure. |
| Peer Network | Patient and caregiver communities that exchange experiences, treatment information, and healthcare navigation advice. |
| ***Clinical facilitators*** |  |
| Clinician Expertise | Advanced knowledge, skills, and judgement of treating clinicians that supported diagnostic clarification or treatment outcomes. |
| Specialist Input | Additional insights from subspecialists or senior consultants through formal consultations or second opinions. |
| Specialized Interventions | Advanced diagnostic tests or treatments such as PET-CT, genomic testing, novel drugs, clinical trials, or CAR-T therapy. |
| ***Patient-level facilitators*** |  |
| Patient Self-Advocacy | Active involvement in seeking information, contacting specialists, requesting further assessment, or evaluating treatment options. |
| Family Support | Emotional, practical, or financial assistance from relatives that facilitates diagnosis and treatment. |

### Multivariable Regression Analysis

Collinearity testing found weak pairwise correlations, except between tertiary hospitals and peer networks (Cramér’s V=0.44), and between specialized interventions and specialist input (Cramér’s V=0.40), but both pairs were retained given their distinct clinical implications.

In the primary model, all barriers were significantly associated with higher odds of severe-burden status (Figure 3, Table S2). Prior inappropriate treatment showed the strongest association (OR 37.76, 95% CI 21.59-66.04), followed by system-level inefficiency (OR 7.61, 5.33-10.89), and patient-level barriers (OR 4.94, 2.89-8.44). Among facilitators of diagnostic revision, clinician expertise, specialist input, peer network engagement, and specialized interventions were significantly associated with lower odds of severe-burden status (ORs ranging from 0.34 to 0.54). In the sensitivity analysis excluding prior inappropriate treatment, all associations remained directionally consistent with those of the primary model, although facility resource gaps and tertiary hospital care no longer reached statistical significance (Table S2).

**Figure 3.**
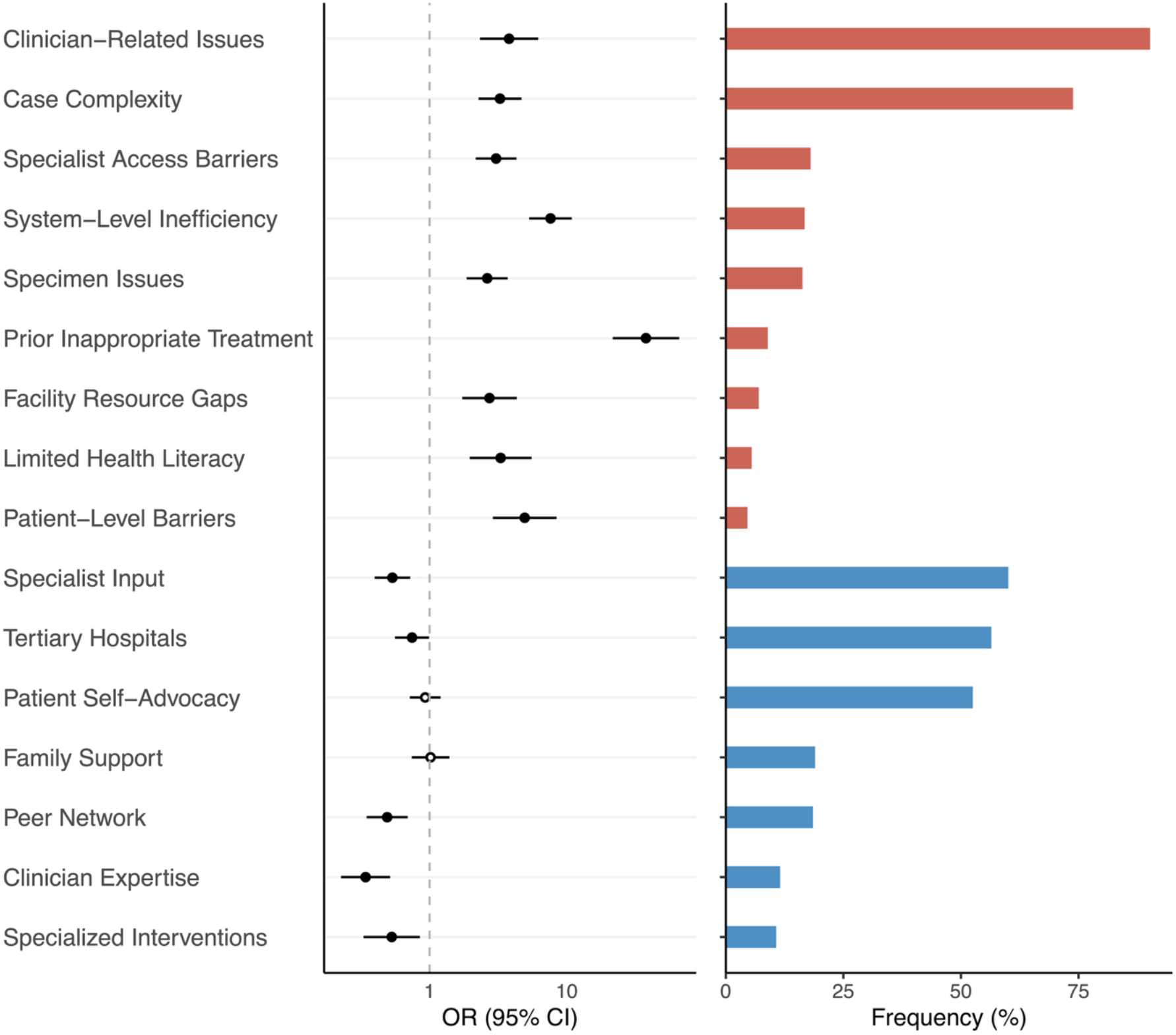
Barriers and facilitators associated with severe-burden status in diagnostic revision. Left: Forest plot from multivariable logistic regression showing odds ratios (ORs) and 95% confidence intervals (CIs) for narrative-derived barriers and facilitators associated with severe-burden status. Filled dots indicate p≤0·05; open dots indicate p>0·05. Patient-level barriers included patient psychological resistance (n=53, 2·9%), family or financial constraints (n=32, 1·8%), and patient physical limitations (n=8, 0·4%). Right: Frequency of barriers and facilitators across the analytic cohort (n=1,801). Bars represent the proportion of narratives mentioning each factor, with barriers and facilitators ordered by descending frequency within each category. Both panels share the same ordering.

Human audit yielded a PPV of 90.6% for identifying eligible cases and a human-AI agreement (κ) of 0.64 for burden status classification. In a supplementary audit of 165 cases flagged by our pipeline as involving prior inappropriate treatment, three recurring patterns were identified: (i) initial lymphoma subtype misclassification leading to treatment regimens later revised after specialist review (n=34, 20.6%); (ii) initial attribution to inflammatory or infectious causes resulting in prolonged ineffective therapy (n=48, 29.1%); and (iii) initial misclassification as a solid tumour, leading to surgical procedures later deemed unnecessary after pathology confirmation (n=57, 34.5%).

## DISCUSSION

### Key Findings

This large-scale AI-assisted study of online forum narratives identified eleven barriers and seven facilitators related to lymphoma diagnostic revision. Among barriers, clinician-related issues and case complexity were most frequent, yet prior inappropriate treatment, with relatively low frequency, showed the strongest association with severe-burden status. Among facilitators, specialist input, tertiary hospital care, and patient self-advocacy were most frequently reported, yet clinician expertise and peer networks showed the strongest association with lower odds of severe-burden status. Together, these results characterised patient experiences with lymphoma diagnostic revision as a pathway shaped by barriers, facilitators, and varying levels of associated burden.

### Comparison with Existing Evidence

Our findings aligned with prior work on cancer care quality, where delays, fragmentation, and inadequate psychosocial support were key problems faced by patients, and multidisciplinary coordination acted as facilitators.^12^ Moreover, we found case complexity (74.1%) and specimen issues (16.5%) to be significant barriers, consistent with long-standing recognition that pathology and histopathological interpretation in lymphoma are inherently challenging.^2-4^

The recent GPS on lymphoma and chronic lymphocytic leukaemia revealed significant information gaps and treatment regret, with Chinese respondents reporting particularly severe challenges: only 15-18% understood their diagnostic process (vs 59% globally), and 53% expressed treatment regret (vs 11% globally).^10,24^ However, that survey did not examine specific risk factors or mechanisms underlying these disparities. Our study extends this literature by providing a risk prioritisation framework derived directly from online patient experiences in a Chinese context, distinguishing high-frequency issues from high-risk events. For example, the “information gaps” highlighted in the GPS may relate in our data to limited health literacy (5.7% of cases) and clinician-related issues (90.4%). While the high frequency of clinician-related issues does not necessarily imply malpractice, it reveals patient-perceived gaps and communication breakdowns that compromise quality of care.^6,25^ “Treatment regret” in the GPS may also partly reflect patient experiences related to inappropriate prior treatment in our dataset, which was reported in only 9.2% of cases but manifested as the strongest predictor (OR 37.76), exemplifying the risks of initiating therapy without diagnostic certainty. Beyond individual clinician decisions, systemic factors including system-level inefficiency, specialist access barriers, and facility resource gaps were also associated with severe burdens. Patient and family-level barriers posed substantial, though less frequent risks.

Among the facilitators, clinician expertise, specialist input and specialized interventions represented positive forces from within the formal healthcare system. Outside the system, peer networks, formed by patients and caregivers, emerged as an important source of information and support, in contrast to patient self-advocacy and family support, which showed no significant association with burden severity. This pattern suggests that individual-level reactive efforts may be insufficient in isolation, whereas collective patient networks may function as proactive intermediaries capable of mitigating diagnostic risk. Functionally, advice from highly knowledgeable patient advocates often served roles similar to those of healthcare professionals, providing timely interpretation and referral guidance when access to formal care was fragmented. Cancer patients have been shown to actively seek peer stories to better understand treatment choices, manage emotional impact, and reduce feelings of isolation.^13^ Such experience-sharing may complement clinical information and foster emotional coping, particularly in the face of diagnostic uncertainty.

### Strengths and Limitations

To our knowledge, this is the first large-scale, AI-assisted analysis of patient experiences with diagnostic revision in lymphoma. We developed a scalable, reproducible, and potentially adaptable AI pipeline to transform heterogeneous online forum narratives into structured case profiles. Our function-based prompt design formalised diagnostic revision from natural language, which helped reduce semantic ambiguity in LLM prompting and improve case extraction accuracy. Transformer-assisted keyword clustering enabled bottom-up identification of barriers and facilitators of diagnostic revision, reducing reliance on pre-defined assumptions. In addition, the human audit framework evaluated LLM-generated outputs against an explicit reverse-engineered scoring rubric, offering transparency into how LLM interpreted inherently subjective patient experiences. This collaborative process aligned with emerging efforts to integrate AI with human judgement in complex, subjective content analysis tasks.^21,22^

Several limitations should be noted. First, this study focused on a single Chinese-language platform, with users potentially representing a more activated, health-literate subset of patients. Second, all findings are based on retrospective narratives with possible recall bias and subjective framing. Diagnostic revision itself was identified from forum narratives, with uploaded clinical materials where available, but was not independently verified against medical records. Discussion of prior inappropriate treatment may trigger reinterpretation of earlier experiences, inflating perceived treatment-related harm or burden severity. A sensitivity analysis excluding this barrier confirmed directional consistency for other factors, supporting the robustness of the overall pattern. Nonetheless, the unusually large association may be inflated due to potential criterion contamination between the predictor and the outcome, rather than a generalisable estimate across broader diagnostic contexts. Third, we did not map LLM-extracted diagnoses to International Classification of Diseases codes (ICD-9/ICD-10) for standardised classification, nor perform external validation of the AI-extracted fields through forum user engagement, which could otherwise have enhanced credibility.Fourth, the anonymous nature of online platforms, along with OCR limitations and LLM input constraints, may have led to information loss. Salience-based keyword extraction may also have led to under-ascertainment of less prominent factors. Fifth, given the observational design, regression associations and temporal trends should be interpreted as correlations rather than causal effects. Finally, online forum narratives cannot fully disentangle diagnostic errors from appropriate and necessary diagnostic evolution in complex diseases such as lymphoma.

### Implications for Diagnostic Safety and Digital Health

Health systems often prioritize frequent problems, but patient narratives reveal that the most harmful risks may arise from relatively uncommon but high-impact failures. For patient-centred lymphoma care, digital health tools may offer solutions across multiple levels. At the clinical level, AI-assisted pathology systems have shown strong performance in lymphoma detection and may reduce diagnostic complexity and specialist workload.^7,8^ At the system level, digital platforms including decision-support tools, triage algorithms, and telemedicine could alleviate financial, logistical, and physical constraints in under-resourced settings. At the patient level, LLM applications such as conversational agents may enhance health literacy, provide psychosocial support, and facilitate shared decision-making, potentially mitigating clinician-patient friction and improving understanding of the diagnostic process.^25-27^

Notably, some patients in our sample reported using LLMs to interpret pathology reports or clarify medical uncertainties. This pattern may be partly enabled by China’s healthcare context, where patients and family caregivers often have ready access to personal clinical documents and test reports, allowing them to seek, interpret, and verify medical information beyond individual clinical encounters. While prior studies have cautioned against over-reliance on AI-generated advice, ^28^ our findings point to a parallel reality where patients turned to AI for its perceived neutrality and accessibility in response to systemic failures, diagnostic uncertainty, or fragmented care. This underscores the need for health systems to integrate safe, patient-facing AI tools into routine care with appropriate oversight. More broadly, digital platforms now enable patients to self-organize, form peer networks, exchange information, and seek emotional support. These online communities serve as informal knowledge intermediaries that complement formal care and signal unmet needs. Health systems may benefit from recognizing these ecosystems as valid windows into patient perspectives. Integrating insights from patient-generated content could inform service design, policy reform, and communication strategies to improve quality of care and patient experience.

### Future Research

Future work may explore longitudinal analysis of patient narratives to understand how barriers and facilitators evolve over time and to distinguish short-from long-term burdens. Cross-system comparison of patient-reported experiences with diagnostic revision is feasible by adapting this pipeline to comparable patient communities in other social platforms and languages. Beyond lymphoma forum data, our analytic framework could be applied to other forms of clinical or narrative data across diseases and healthcare settings, enabling systematic incorporation of patient perspectives into clinical quality assessment and digital health innovation.

## CONCLUSION

This study of 1,801 cases from online forum narratives identified eleven barriers and seven facilitators along lymphoma diagnostic revision pathways. Our findings demonstrate that diagnostic safety requires not only hospital-based interventions but also coordinated efforts involving patients, families, peer networks, and system-level infrastructure. More broadly, this study shows the feasibility of leveraging AI to analyse patient-generated data at scale, offering a novel lens to inform clinical practice, health policy, and digital health innovation.

## Data Availability

The de-identified dataset may be shared upon reasonable request.

## CONFLICT OF INTEREST

All authors declare no competing interests.

## AUTHOR CONTRIBUTIONS

FH: Conceptualization, methodology, software, data curation, formal analysis, visualization, writing (original draft), project administration. ZY: Methodology, validation, writing (review and editing). XZ: Methodology, validation, writing (review and editing). ZY and XZ contributed equally to this work. WLK: Validation, writing (review and editing). LLS: Validation, writing (review and editing). NG: Methodology, writing (review and editing).JMV: Conceptualization, supervision, methodology, writing (review and editing).

## ACKNOWLEDGMENTS

We thank participants of online patient forums for sharing their experiences, which made this research possible. ChatGPT-5.3, Claude Sonnet 4.5, and Claude Opus 4.6 were used for language editing, and to assist with code development. All scientific content and analyses were conducted and verified by the authors.

## DATA & CODE AVAILABILITY

All codes and study protocols used for data collection, processing, audit, and statistical analysis will be made publicly available upon publication. Aggregated data are available from the corresponding author upon reasonable request, subject to institutional and ethical approvals.

## Notes

### Competing Interest Statement

The authors have declared no competing interest.

### Funding Statement

This study did not receive any funding

### Author Declarations

The study used ONLY openly available human data that were originally located at House086.com.

### Summary of Updates

Substantial revisions have been made since the previous preprint version. Key changes include: 1. Updated the schema for LLM-assisted case extraction. 2. Added a rubric-guided human audit. 3. Updated the author list to reflect additional contributions. 4. Refined the manuscript framing and revised the figures, tables, and supplementary materials accordingly. 5. Updated the funding statement.

## REFERENCES

1. Sung H, Ferlay J, Siegel RL, et al. Global cancer statistics 2020: GLOBOCAN estimates of incidence and mortality worldwide for 36 cancers in 185 countries. CA: a cancer journal for clinicians 2021; 71(3): 209–49.

2. Proctor IE, McNamara C, Rodriguez-Justo M, Isaacson PG, Ramsay A. Importance of expert central review in the diagnosis of lymphoid malignancies in a regional cancer network. Journal of Clinical Oncology 2011; 29(11): 1431–5.

3. Bowen JM, Perry AM, Laurini JA, et al. Lymphoma diagnosis at an academic centre: rate of revision and impact on patient care. Br J Haematol 2014; 166(2): 202–8.

4. Laurent C, Baron M, Amara N, et al. Impact of expert pathologic review of lymphoma diagnosis: study of patients from the French Lymphopath Network. Journal of clinical Oncology 2017; 35(18): 2008–17.

5. Deng J, Zuo X, Yang L, Gao Z, Zhou C, Guo L. Misdiagnosis analysis of 2291 cases of haematolymphoid neoplasms. Frontiers in Oncology 2023; 13: 1128636.

6. Singh H, Schiff GD, Graber ML, Onakpoya I, Thompson MJ. The global burden of diagnostic errors in primary care. BMJ Qual Saf 2017; 26(6): 484–94.

7. Syrykh C, Abreu A, Amara N, et al. Accurate diagnosis of lymphoma on whole-slide histopathology images using deep learning. NPJ digital medicine 2020; 3(1): 63.

8. Li D, Bledsoe JR, Zeng Y, et al. A deep learning diagnostic platform for diffuse large B-cell lymphoma with high accuracy across multiple hospitals. Nature communications 2020; 11(1): 6004.

9. Mazor KM, Roblin DW, Greene SM, et al. Toward patient-centered cancer care: patient perceptions of problematic events, impact, and response. Journal of Clinical Oncology 2012; 30(15): 1784–90.

10. Lymphoma Coalition. 2024 Global Patient Survey on Lymphomas & CLL. 2024. https://lymphomacoalition.org/global-patient-survey/ (accessed Jan 04, 2026).

11. Tsianakas V, Maben J, Wiseman T, et al. Using patients’ experiences to identify priorities for quality improvement in breast cancer care: patient narratives, surveys or both? BMC Health Services Research 2012; 12(1): 271.

12. Wagner EH, Aiello Bowles EJ, Greene SM, et al. The quality of cancer patient experience: perspectives of patients, family members, providers and experts. Quality and Safety in Health Care 2010; 19(6): 484.

13. Engler J, Adami S, Adam Y, et al. Using others’ experiences. Cancer patients’ expectations and navigation of a website providing narratives on prostate, breast and colorectal cancer. Patient Education and Counseling 2016; 99(8): 1325–32.

14. Ning K, Gu H, Franklin M, et al. Online Health-Seeking Behaviors and Information Needs Among Patients With Lymphoma in China: Study of Regional and Temporal Trends. Journal of Medical Internet Research 2025; 27: e80497.

15. Dreisbach C, Koleck TA, Bourne PE, Bakken S. A systematic review of natural language processing and text mining of symptoms from electronic patient-authored text data. Int J Med Inform 2019; 125: 37–46.

16. Siddiqui ZA, Pathan M, Nduaguba S, et al. Leveraging social media data to study disease and treatment characteristics of Hodgkin’s lymphoma Using Natural Language Processing methods. PLOS Digit Health 2025; 4(3): e0000765.

17. Hsieh H-F, Shannon SE. Three approaches to qualitative content analysis. Qualitative health research 2005; 15(9): 1277–88.

18. Elo S, Kyngäs H. The qualitative content analysis process. Journal of advanced nursing 2008; 62(1): 107–15.

19. JaidedAI. EasyOCR: Ready-to-use Optical Character Recognition with 80+ Supported Languages. 2020. https://github.com/JaidedAI/EasyOCR (accessed Sep 2, 2025).

20. Gibney E. China’s cheap, open AI model DeepSeek thrills scientists. Nature 2025; 638(8049): 13–4.

21. Yang M, Luo Y, He T, et al. Application of artificial intelligence to measure and predict patient values and preferences: a scoping review. npj Digital Medicine 2025; 8(1): 769.

22. Vaccaro M, Almaatouq A, Malone T. When combinations of humans and AI are useful: A systematic review and meta-analysis. Nature Human Behaviour 2024; 8(12): 2293–303.

23. Rea LM, Parker RA. Designing and conducting survey research: A comprehensive guide: John Wiley & Sons; 2014.

24. House086. Insights of GPS Survey | The Dual Burdens and Support Deficits Faced by Lymphoma Families in China. 2025. https://www.house086.com/thread-330403-1-1.html (accessed Jan 08, 2026).

25. Salwei ME, Ancker JS, Weinger MB. The decision aid is the easy part: workflow challenges of shared decision making in cancer care. JNCI: Journal of the National Cancer Institute 2023; 115(11): 1271–7.

26. Thirunavukarasu AJ, Ting DSJ, Elangovan K, Gutierrez L, Tan TF, Ting DSW. Large language models in medicine. Nature Medicine 2023; 29(8): 1930–40.

27. Arora A, Arora A. The promise of large language models in health care. The Lancet 2023; 401(10377): 641.

28. Shekar S, Pataranutaporn P, Sarabu C, Cecchi GA, Maes P. People Overtrust AI-Generated Medical Advice despite Low Accuracy. NEJM AI 2025; 2(6): AIoa2300015.

