## Supplementary for "Patient Experiences with Diagnostic Revision in Lymphoma: Analysis of Chinese Online Forum Narratives Using Natural Language Processing"

### SUPPLEMENTARY MATERIALS

#### Method S1. Prompt logic for detecting diagnostic revision from forum narratives.

To minimize the ambiguity of natural languages, we defined diagnostic revision as a mapping function $f(X)=Y$, from an initial diagnosis $X$ to a revised diagnosis $Y$ $(Y \neq X)$. The function-based prompt instructed the DeepSeek model to analyse discussion threads from the House086 lymphoma forum and identify all cases satisfying at least one of the following criteria:

1. Explicit diagnostic chain: The narrative explicitly describes two or more distinct or conflicting diagnoses (e.g., initially diagnosed as $X$, later as $Y$, with $X \neq Y$).

2. Implicit diagnostic chain: The patient was initially suspected or treated for $X$, but later confirmed as $Y$ through pathological reassessment.

3. Third-party case description: The narrative provides another specific patient, with sufficient details to re-construct a diagnostic trajectory from $X$ to $Y$.

4. Keyword-based cues (borderline cases to be manually reviewed): Narratives or comments explicitly concluded the experience as “misdiagnosis”, “pathology error”, or “revised diagnosis”, even in the absence of full diagnostic chronology.

Included cases typically involved: $X$= benign conditions, $Y$=lymphoma; $X$= solid tumours, $Y$= lymphoma; and subtype reclassifications within lymphoma. We excluded cases with incomplete revisions (missing $Y$), unsubstantiated suspicions, unchanged diagnoses, or diagnostic shifts attributable to disease progression rather than error.

The full prompt specification and extraction schema are provided in a publicly available repository [to be released upon publication].

### Method S2. Human audit grading rubric for burden severity, translated from Chinese.

**Evaluation Core**: Rooted in the patient's perspective and experience, and synthesized with medical logic, these criteria assess the cumulative and holistic burden resulting from misdiagnosis, delays, or inappropriate clinical interventions during the "diagnostic revision" process. The evaluation prioritizes the severity and reversibility of physiological harm, long-term health risks, loss of therapeutic opportunities, and the impact on the patient's baseline physiological/psychological status at the time correct treatment is initiated.

**5 = Catastrophic**: Directly results in patient death, permanent severe disability, or the irreversible loss of vital organ function.

**4 = Severe**: Significantly alters the patient's lifestyle, survival expectation, or physiological functional reserve. These cases involve a high probability of increased difficulty and risk in subsequent treatments or have already caused long-term, substantial, and potentially partially irreversible harm. Level 4 is distinguished from Level 3 by the presence of profound, long-term impact and the deprivation of optimal therapeutic windows. Examples include:

- Inappropriate Treatment Harm: Exposure to high-risk or high-toxicity treatments mismatched with the final diagnosis (e.g., unnecessary chemotherapy for a benign condition), leading to the loss of first-line treatment opportunities or causing severe complications and long-term toxicities.
- Invasive Intervention Harm: Undergoing avoidable, organ-damaging major surgery that carries high risks of complications, exceeds the necessary costs of diagnosis/emergency care, or results in permanent partial loss of organ function.
- Consequences of Delay: Depletion of physiological reserves or significant disease progression due to delays, which substantially increases treatment complexity and worsens prognosis.
- Holistic Socio-economic Harm: Severe life disruptions such as family breakdown, catastrophic debt, or withdrawal from education, which significantly lower future life expectations.
- Extreme Suffering: Prolonged periods of extreme physical pain or psychological distress directly caused by misdiagnosis or delay.
- Institutional Trust Crisis: A breakdown of trust caused by a physician's stubborn adherence to an erroneous diagnosis, forcing the patient to navigate multiple institutions and resulting in major psychological trauma that hinders subsequent treatment adherence.

**3 = Moderate**: Short-term harm that is recoverable or compensable through proper clinical management, or represents manageable and necessary costs incurred during the diagnostic process. Recovery does not alter long-term lifestyle or quality of life, and the harm does not interfere with the timely initiation or efficacy of subsequent standard treatment. Level 3 is distinguished from Level 4 by the absence of long-term risks or elevation of the therapeutic threshold. Examples include:

- Repeated invasive diagnostic procedures (e.g., multiple biopsies due to suboptimal sampling) causing transient pain without long-term sequelae.
- Initial treatments with low toxicity (e.g., anti-inflammatory injections, herbal medicine) that were non-standard or unnecessary but lacked long-term side effects and did not delay core treatment.
- Necessary Diagnostic/Emergency Costs: Management of acute complications not caused by delay (e.g., emergency resection for bowel obstruction) or invasive procedures required by difficult anatomical locations, provided the intervention was clinically indicated and the patient recovered well.
- Short-to-medium-term physical or psychological distress where the disease remained stable and symptoms resolved fully following the correct diagnosis.
- Proactive Systemic Correction: Scenarios where a physician proposed high-risk erroneous treatments but corrected the course before implementation, preventing substantive harm and maintaining the patient-provider trust.

**2 = Mild**: Distinguished from Levels 3–5 by the fact that the patient has not yet received any pharmacological or therapeutic treatment (including anti-inflammatories or herbal medicine) and the disease has not progressed.

- Undergoing minimally invasive examinations (e.g., superficial fine-needle aspiration).
- Logistical burdens, such as repeated outpatient visits, resulting in significant time loss, work absence, or minor financial expenditure.
- Short-term anxiety regarding diagnostic uncertainty without substantive physiological harm.

**1 = None / Minimal**: The diagnostic error is corrected rapidly without financial loss, invasive procedures, incorrect treatment, or detectable disease progression.

**Table S1. Keywords used to identify diagnostic revision threads**

| Category | Keywords |
|---|---|
| Direct terms | 会诊 (second opinion), 误诊 (misdiagnosis), 漏诊 (missed diagnosis), 异议 (discrepancy), 争议(conflict), 不一样 (inconsistency) |
| Indirect terms | 延误 (delay), 错误 (error), 换医院 (changing hospital), 责任 (accountability), 病理 (pathology) |
| Pathology experts | Names of leading national lymphoma pathology experts (used as proxies for second-opinion seeking. |
| Frequently reported initial diagnoses | 乳腺癌 (breast cancer), 肺癌 (lung cancer), 甲状腺癌 (thyroid cancer), 胃癌 (gastric / stomach cancer), 胸腺瘤 (thymoma), 炎症 (inflammation), 结核 (tuberculosis), 腮腺炎 (parotitis / mumps), 阑尾炎 (appendicitis) |
| Other expressions | 手术 (surgery), 术后(post-surgery), 怀疑 (suspicion), 消炎 (anti-inflammatory / antibiotic), 中药 (traditional Chinese medicine) |

**Table S2. Regression results for barriers and facilitators associated with severe-burden status**

| | Frequency | Main Analysis | | Sensitivity Analysis | |
|---|---|---|---|---|---|
| | Count (%) | OR [95% CI] | p-value | OR [95% CI] | p-value |
| **Barriers** | | | | | |
| Clinician-Related Issues | 1629 (90.4) | 3.80 [2.33 - 6.20] | <0.0001 | 3.19 [2.02 - 5.02] | <0.0001 |
| Case Complexity | 1334 (74.1) | 3.26 [2.27 - 4.68] | <0.0001 | 1.48 [1.09 - 2.01] | 0.013 |
| Specialist Access Barriers | 329 (18.3) | 3.05 [2.17 - 4.30] | <0.0001 | 1.40 [1.03 - 1.90] | 0.032 |
| System-Level Inefficiency | 306 (17.0) | 7.61 [5.33 - 10.89] | <0.0001 | 3.12 [2.29 - 4.27] | <0.0001 |
| Specimen Issues | 298 (16.5) | 2.63 [1.86 - 3.72] | <0.0001 | 1.35 [0.98 - 1.84] | 0.062 |
| Prior Inappropriate Treatment | 165 (9.2) | 37.76 [21.59 - 66.04] | <0.0001 | NA | NA |
| Facility Resource Gaps | 130 (7.2) | 2.73 [1.73 - 4.33] | <0.0001 | 1.30 [0.85 - 1.98] | 0.22 |
| Limited Health Literacy | 103 (5.7) | 3.30 [1.96 - 5.55] | <0.0001 | 1.67 [1.02 - 2.71] | 0.040 |
| Patient-Level Barriers* | 87 (4.8) | 4.94 [2.89 - 8.44] | <0.0001 | 2.21 [1.34 - 3.64] | 0.0019 |
| **Facilitators** | | | | | |
| Specialist Input | 1086 (60.3) | 0.54 [0.40 - 0.72] | <0.0001 | 0.65 [0.5 - 0.86] | 0.0022 |
| Tertiary Hospitals | 1021 (56.7) | 0.75 [0.56 - 0.99] | 0.045 | 0.84 [0.65 - 1.09] | 0.20 |
| Patient Self-Advocacy | 950 (52.7) | 0.93 [0.72 - 1.20] | 0.58 | 1.07 [0.84 - 1.35] | 0.59 |
| Family Support | 346 (19.2) | 1.02 [0.74 - 1.40] | 0.92 | 1.17 [0.88 - 1.57] | 0.28 |
| Peer Network | 338 (18.8) | 0.49 [0.35 - 0.69] | <0.0001 | 0.61 [0.44 - 0.83] | 0.0020 |
| Clinician Expertise | 212 (11.8) | 0.34 [0.23 - 0.52] | <0.0001 | 0.49 [0.34 - 0.70] | 0.00013 |
| Specialized Interventions | 197 (10.9) | 0.53 [0.33 - 0.85] | 0.0086 | 0.59 [0.38 - 0.89] | 0.013 |

Outcome: severe-burden status, defined as a DeepSeek-assigned burden severity score $\geq 4$. Multivariable logistic regression adjusted for all barriers and facilitators, additionally adjusted for patient age, patient sex, narrator type, year of posting, and the number of diagnostic events. All continuous variables were converted to categorical variables to accommodate missing values. In sensitivity analysis, prior inappropriate treatment was excluded from the regression formula.

Abbreviations: OR, odds ratio; CI, confidence interval; NA, not available.

*Patient-level barriers: patient psychological resistance (n=53, 2.9%), family or financial constraints (n=32, 1.8%), or patient physical limitations (n=8, 0.4%).
